# Predictors of gestational weight gain and its association with post-natal growth of children under two years

**DOI:** 10.1101/2025.05.14.25327642

**Authors:** Mahama Saaka, Abel Mahami Dauda

## Abstract

**Background:** Although majority of previous studies have investigated the relationship between gestational weight gain (GWG) and pregnancy outcomes, there is a paucity of information on how GWG relates with post-natal growth outcomes. This study assessed the predictors of GWG and how it associates with postnatal growth outcomes of children under two years in the East Mamprusi Municipality of Ghana.

**Methods:** A facility-based analytical cross-sectional study was conducted among 320 mother-child pairs attending child welfare clinics in selected health facilities. Binary logistic regression was used to assess the factors associated with inadequate gestational weight gain and multivariable linear regression was used to identify predictors of length-for-age z scores (LAZ).

**Results:** Inadequate GWG rate was high in the population at 90.3% based on the recommended IOM criteria on GWG. The key predictors of GWG were first trimester BMI and gravidity. A unit increase in the first trimester BMI was associated with 31 % protection against inadequate rate of GWG, AOR= 0.69 (95% CI: 0.58 to 0.82, and p<0.001). Children of women with inadequate GWG had a significant lower LAZ, compared with their colleagues who were born to women who had adequate GWG [beta coefficient (β), = -0.181 (95% CI: -2.72 to -0.14, p = 0.03)]. Inadequate GWG was associated with low length-for-age z scores (LAZ) among children aged 0-6 months.

**Conclusions:** There is a need for targeted nutritional programs to support adequate weight gain during pregnancy, especially for mothers with higher gravidity so as to reduce the prevalence of child stunting.

## Introduction

In most low- and middle-income countries (LMICs), including Ghana, inadequate gestational weight gain (GWG) has been reported to be a major public health concern [1, 2] and it is associated with an increased risk of low birth weight (LBW) and pre-term birth [3-7]. GWG is a potentially modifiable factor and it is an important indicator of maternal and foetal nutrition [8-10]. It is well documented that child growth failure occurs from conception up-to two years of age, and about 50% of the growth failure which occurs by two years of age takes place during the gestation period [11-14]. This may be attributed to the established understanding that an insult which occurs during pregnancy permanently affects tissue structure and function [13]. Inadequate GWG, increases the risk of intrauterine growth restriction (IUGR), irrespective of pre-pregnancy body mass index (BMI), [15-17].

The majority of previous studies in the literature have focused on the association between weight gain during pregnancy and outcomes at birth [6, 7, 18-21]. Little is, however, documented on how GWG in pregnancy relates with postnatal growth outcomes. Furthermore, knowing the factors that affect GWG will help in taking appropriate nutrition-related behavioral measures. Therefore, this study assessed the predictors of GWG and how it is associated with postnatal growth outcomes of children under two years in a malaria-endemic setting.

## Key messages

- Although majority of previous studies have investigated the relationship between gestational weight gain (GWG) and pregnancy outcomes, there is a paucity of information on how GWG relates with post-natal growth outcomes
- The results of the current study showed that inadequate GWG was associated with low length-for-age z scores (LAZ) among children aged 0-6 months.
- The association between GWG and postnatal growth outcomes suggests that targeted interventions are needed to address maternal nutrition, particularly among multigravida women and those with lower pre-pregnancy

## Materials and Methods

### Study setting and participants

The study was undertaken in the East Mamprusi Municipal District in the North East Region of Ghana. The district has a total population of about 188,006 as of 2021 with 51.5% of the population being females and 48.5% males [22]. Agriculture employs about 90% of the population in the Municipality. Food Crops cultivated include maize, rice and cassava. The health services in the municipality cover broad areas including disease surveillance, nutrition, reproductive and child health, clinical and support services.

The study population comprised 320 mother-child pairs who were attending child welfare clinics (CWC) in some selected facilities in the study area. All mothers who attended antenatal care (ANC) clinic during the first trimester of their last pregnancy and gave birth to a live baby in the same health care facilities were recruited for the study. An additional criterion for recruitment was that the child should be under the age of two years. The exclusion criteria were as follows: incomplete data for the mother and her offspring during pregnancy, multiple pregnancies, preeclampsia/ eclampsia, offspring congenital disease, mothers who initiated ANC in the second or third trimester of their pregnancies.

### Study design, sample size calculation and sampling

A facility-based, analytical cross-sectional study design was used. A two-stage sampling technique involving simple random and systematic sampling were employed in selecting the study participants. Simple random sampling method was used in the selection of five study health facilities out of seven health facilities. At the health facilities, systematic random sampling was applied to select the study subjects. The sampling frame was the list of women contained in the postnatal care (PNC) register. On each PNC day, the mothers who met the inclusion criteria were selected systematically. This was done until the desired sample size was achieved.

The minimum sample size was estimated based on a single population proportion formula with the assumption of 95% confidence level and 0.05 margin of error. The prevalence of stunted child growth, which is the main outcome of interest was estimated to be 29.6 % among children in the North East Region [22] and so the minimum sample size was estimated to be 320.

### Data collection tools and measurements

The primary data was collected through face-to-face interviews using structured questionnaires and checklists. Primary data collected included sociodemographic characteristics, dietary diversity of children, child anthropometric measurements. Secondary data including maternal weight and height, gestational age, number of ANC visits, gravidity and parity were extracted from a review of maternal and child health records books (MCHRB). The questionnaire was hosted on Kobo Collect software to collect the data electronically and transported to MS-Excel software for cleaning. **Recruitment of study participants started on 10/05/2024 and ended 05/10/2024**.

### Study variables and measurement

The main exposure variable of this study was gestational weight gain (GWG). The covariate variables included maternal factors, such as age, pre-pregnancy BMI, last delivery mode, lifestyle before and during pregnancy, education level, parity, employment status, marital status dietary patterns, prenatal care attendance etc. The main outcome measures were postnatal childhood growth indicators including stunting, which was defined as a height-for-age z score less than -2. The key study variables were measured as shown below:

### Assessment of gestational weight gain

GWG was determined as the difference between the weight of each woman in the third trimester (that is, approximately 36 weeks gestation) and weight in the first trimester (that is, in the first 12 weeks of gestation). The rate of GWG per week was calculated by dividing the total weight gain in grams by the gestational ages in weeks, making it independent of the pregnancy duration. Weekly GWG rate was categorized as inadequate, adequate, or excessive, in relation with first trimester-pregnancy BMI and according to the revised 2009 American Institute of Medicine (IOM) GWG guidelines [23]. The National Academy of Medicine, formerly the Institute of Medicine (IOM), recommended GWG rate ranges are: (0.44– 0.58 kg/week for underweight, 0.35–0.50 kg/week for normal weight, 0.23–0.33 kg/week for overweight, and 0.17–0.27 kg/week for obese). GWG was categorized as inadequate (if weekly rate of GWG was below the recommended range), adequate (if weekly rate of GWG was within the recommended range), or excessive (if weekly rate of GWG was above the recommended range) [23].

### Postnatal child growth assessment

The primary outcome of interest was post-natal child growth as measured by length-for-age z-score (LAZ) and weight-for-length z-score (WLZ), and weight-for-age z-score (WAZ) and these were calculated as recommended by the World Health Organization [24]. The continuous z-scores were also categorized as stunting (LAZ < –2 SD), wasting (WLZ < –2 SD) and underweight (WAZ< –2 SD) from the median of the reference population.

### Assessment of child and maternal anthropometry

The mother’s height was measured using a height measure to the nearest 0.1 cm. Weight was measured without shoes and heavy clothing to the nearest 100 g. Body mass index (BMI) was calculated by dividing the body mass in (kilograms) by the square of the body height (in meters). Maternal BMI in early pregnancy (not more than 12 weeks gestation) was also calculated as a proxy for pre-pregnancy BMI [25, 26]. The maternal BMI in early pregnancy was categorized according to the WHO cut off points: underweight (<18.5 kg/m^2^), normal weight (18.5–25.0 kg/m^2^), overweight (25.0^+^–30.0 kg/m^2^) and obesity >30 kg/m^2^.

The child’s length was measured to the nearest 0.1 cm in a recumbent (lying) position using a horizontal wooden length board and movable headpiece (infantometer) and weight was measured to the nearest 100 g using an electronic weighing scale (Seca 874). Children were in light clothing appropriate for the measurements.

### Data quality management

Several measures were taken to ensure the accuracy and reliability of the data collected. Before the data collection, a two-day training was held for the data collectors and supervisors. The content of this training included study objectives and methodology, relevance of the study, ethical concern, and techniques of interviews. There was pre-testing of the questionnaire during the training. In all 5% of the study questionnaires were pre-tested and validated with mother-pair child who were not part of the study. Field supervisors checked for completeness and consistency of the completed questionnaire in the field and during data entry to ensure data quality.

### Data processing and statistical analyses

The data were analyzed using SPSS version 24.0 (SSPS Inc., Chicago, IL, USA). Categorical data are presented as percentages with frequency, and continuous data are presented as mean and standard deviation (SD). The number of women that gained excessively during pregnancy were very few. For the purposes of regression analysis, the rate of GWG per week (g) was categorized into two (inadequate and adequate/ excessive). Binary logistic regression was therefore used to assess the factors associated with inadequate gestational weight gain. Adjusted Odds Ratio (AOR) at 95% confidence intervals were used to assess the strength of associations at p-value <0.05 significance level. Multivariable linear regression was performed to identify the independent effect of gestational weight gain per week on LAZ, adjusting for potential confounding factors. The strength of association was measured using beta coefficients, and their 95% confidence intervals (CI). To check for potential multicollinearity, variance inflation factor (VIF) was calculated. However, the range of calculated VIF values for all the independent variables were 1.02 and 1.06. A value of <5, meant that there was no multicollinearity. Z-scores that were outliers according to the WHO criteria [27]: LAZ (-6, +6), WLZ (-5, +5), WAZ (-6, +5), BMI for-age z score >5 or <−5 and were excluded in the data analysis

### Ethics considerations

Ethical clearance for this study was obtained from the University for Development Studies’ Institutional Review Board (UDSIRB) (Reference no. UDS/RB/101/24/). Preceding data collection, informed written consent was obtained from all the participants after being fully educated on the purpose and scope of this study. The written consent document, signed and dated by the participant and researcher, serves as a record of this agreement. In situations, where the participants could not write or read, thump-printed informed consent was obtained.

## Results

### Socio-demographic characteristics of respondents

Table 1 shows the sociodemographic characteristics of study participants whose mean age was 27.5 ± 5.8 years, and 48.1% were between 25 and 34 years of age. The majority (51.9%) were non-office/Self-employed. In terms of education, 27.5 % of them attained at least Senior High School level education whilst 31.9 % had no formal education and most of them (99.7%) were married.

**Table 1:**
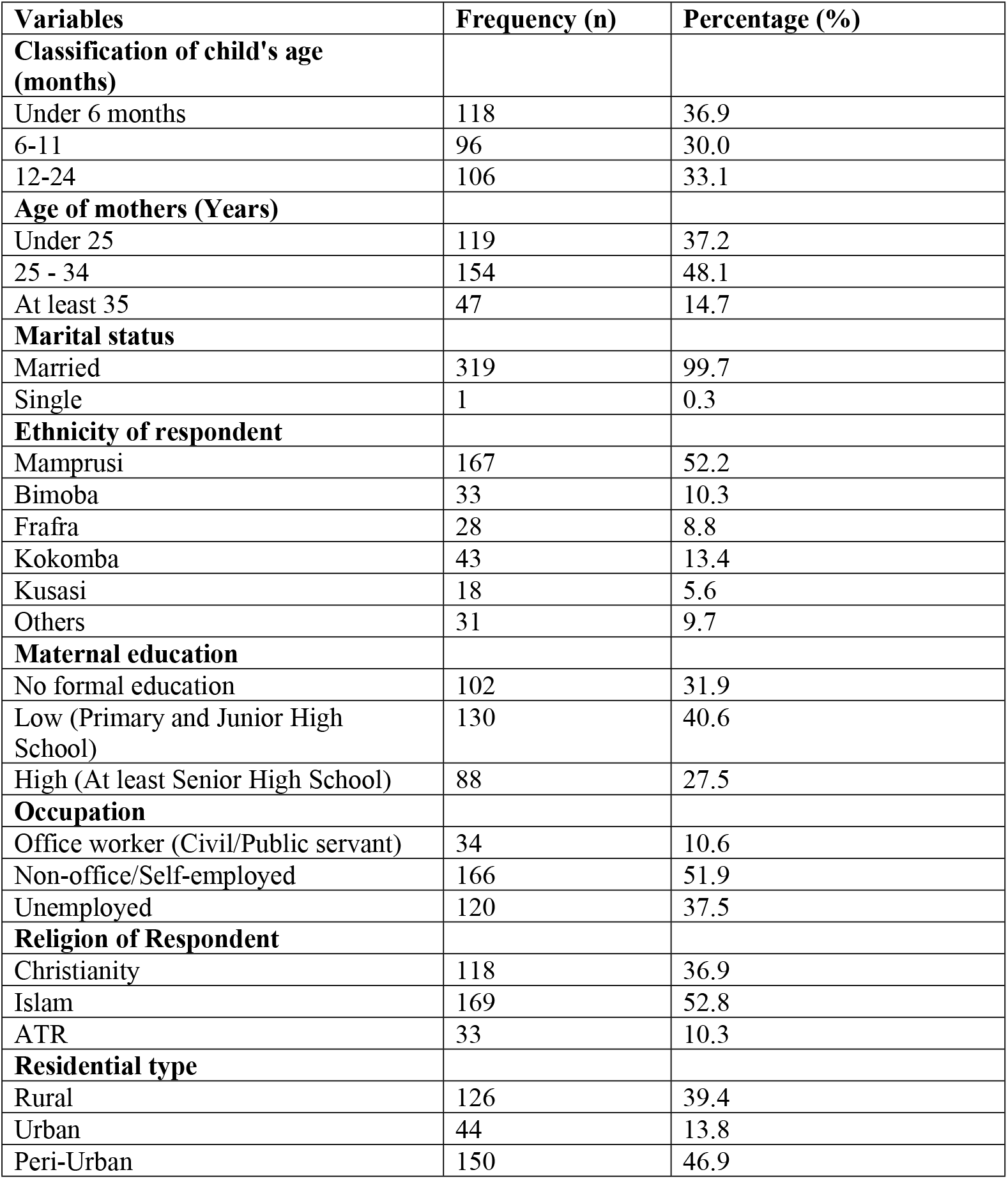
Socio-demographic characteristics of respondents (N = 320)

| **Variables** | **Frequency (n)** | **Percentage (%)** |
| --- | --- | --- |
| **Classification of child's age (months)** |  |  |
| Under 6 months | 118 | 36.9 |
| 6-11 | 96 | 30.0 |
| 12-24 | 106 | 33.1 |
| **Age of mothers (Years)** |  |  |
| Under 25 | 119 | 37.2 |
| 25 - 34 | 154 | 48.1 |
| At least 35 | 47 | 14.7 |
| **Marital status** |  |  |
| Married | 319 | 99.7 |
| Single | 1 | 0.3 |
| **Ethnicity of respondent** |  |  |
| Mamprusi | 167 | 52.2 |
| Bimoba | 33 | 10.3 |
| Frafra | 28 | 8.8 |
| Kokomba | 43 | 13.4 |
| Kusasi | 18 | 5.6 |
| Others | 31 | 9.7 |
| **Maternal education** |  |  |
| No formal education | 102 | 31.9 |
| Low (Primary and Junior High School) | 130 | 40.6 |
| High (At least Senior High School) | 88 | 27.5 |
| **Occupation** |  |  |
| Office worker (Civil/Public servant) | 34 | 10.6 |
| Non-office/Self-employed | 166 | 51.9 |
| Unemployed | 120 | 37.5 |
| **Religion of Respondent** |  |  |
| Christianity | 118 | 36.9 |
| Islam | 169 | 52.8 |
| ATR | 33 | 10.3 |
| **Residential type** |  |  |
| Rural | 126 | 39.4 |
| Urban | 44 | 13.8 |
| Peri-Urban | 150 | 46.9 |

#### 4.3 Medical and obstetric history of mother

Table 2 below presents the obstetric and medical records of the respondents. Less than half (39.4%) of the respondents had malaria infection during pregnancy with index child of which 20 %, and 18.8% had malaria infection once, and at least twice respectively. Majority (53.8 %) of the study participants were multigravida and 71.9 % initiated ANC in the first trimester of pregnancy. The mean first trimester BMI (proxy for pre-pregnancy weight) was 23.67 (SD = 2.80 kg/m^2^) and no mother was underweight (that is, BMI < 18.5). The mean GWG rate was low (162.0±82.5 g/week), with 90.3% of the women gaining less than the recommended American Institute of Medicine (IOM) GWG criteria. The gestational age at which most of the study participants gave birth was full term (≥37 weeks).

**Table 2:** Medical and obstetric history of study participants.

| Variables | Frequency (n) | Percentage (%) |
| --- | --- | --- |
| **Malaria during pregnancy** |  |  |
| No | 194 | 60.6 |
| Yes | 126 | 39.4 |
| **Frequency of malarial infection during pregnancy** |  |  |
| None | 194 | 60.6 |
| Once | 66 | 20.6 |
| At least twice | 60 | 18.8 |
| **Parity** |  |  |
| Primiparous | 81 | 25.3 |
| Secundiparous | 81 | 25.3 |
| Multiparous | 158 | 49.4 |
| **Classification of Gravidity** |  |  |
| Primigravidae | 70 | 21.9 |
| Secundigravidae | 78 | 24.4 |
| Multigravida | 172 | 53.8 |
| **Gestational age at first ANC visit** |  |  |
| First trimester (≤ 12 weeks) | 230 | 71.9 |
| Beyond first trimester (> 12 weeks) | 90 | 28.1 |
| **Frequency of ANC visits before delivery** |  |  |
| Less than 8 | 151 | 47.2 |
| At least 8 | 169 | 52.8 |
| **Adequacy of prenatal care (at least 8 ANC visits & first ANC in first trimester of pregnancy)** |  |  |
| Inadequate | 167 | 52.2 |
| Adequate | 153 | 47.8 |
| **Mode of delivery of current child** |  |  |
| Normal vaginal delivery | 268 | 83.8 |
| Caesarean section | 52 | 16.3 |
| **Gestation at delivery (weeks)** |  |  |
| Preterm (<37 weeks) | 17 | 5.3 |
| Full term (≥37 weeks) | 303 | 94.7 |
| **First trimester BMI (kg/m<sup>2</sup>)** |  |  |
| Underweight (< 18.5) | 0 | 0.0 |
| Normal (18.5-25.0) | 244 | 76.3 |
| Overweight/obese (> 25.0) | 61 | 19.1 |
| **Rate of GWG per week (g)** |  |  |
| Inadequate | 289 | 90.3 |
| Adequate | 23 | 7.2 |
| Excessive | 8 | 2.5 |

### Association between postnatal child growth outcomes and gestational weight gain (kg/week) among infants under 6 months

Analysis of variance (ANOVA) showed that the rate of gestational weight gain (GWG) was associated positively with LAZ for children under 6 months. Children born to women with adequate GWG had significantly higher length-for-age z score than those born to women who had inadequate GWG (Table 3). This association was not observed among older children aged 6-24 months (results not shown),

**Table 3:** Postnatal child growth indicators according to rate of gestational weight gain (kg/week) among infants under 6 months.

| Growth indicator | Rate of GWG |  |  |  | 95% Confidence Interval for Mean |  |  |
| --- | --- | --- | --- | --- | --- | --- | --- |
| Length-for-age z scores (LAZ) |  | N | Mean | Std. Deviation | Lower Bound | Upper Bound | Test statistic |
|  | Inadequate | 104 | -1.05 | 2.56 | -1.55 | -0.55 | F (2, 117) = 3.98, p = 0.02 |
|  | Adequate | 12 | 1.09 | 1.98 | -0.17 | 2.35 |  |
|  | Excessive | 2 | -0.24 | 0.35 | -3.35 | 2.88 |  |
| Weight-for-length z-scores (WLZ) |  |  |  |  |  |  |  |
|  | Inadequate | 104 | -0.63 | 2.68 | -1.15 | -0.11 | F (2, 117) = 2.49, p = 0.09 |
|  | Adequate | 12 | -2.43 | 2.97 | -4.32 | -0.54 |  |
|  | Excessive | 2 | -1.63 | 0.93 | -10.02 | 6.76 |  |
| Weight for age (WAZ) |  |  |  |  |  |  |  |
|  | Inadequate | 104 | -1.41 | 2.06 | -1.81 | -1.01 | F (2, 117) = 0.02, p = 0.98 |
|  | Adequate | 12 | -1.53 | 2.77 | -3.29 | 0.23 |  |
|  | Excessive | 2 | -1.27 | 0.27 | -3.68 | 1.14 |  |

### Predictors of inadequate gestational weight gain per week

The results showed that high first trimester BMI protected against inadequate rate of gestational weight gain (GWG) whilst higher gravidity and male foetus were risk factors (Table 4). A unit increase in first trimester BMI was associated with 31 % protection against inadequate rate of GWG, AOR= 0.69 (95% CI: 0.58 to 0.82, and p<0.001). A unit increase in gravidity had increased odds of inadequate GWG rate, AOR= 2.39 (95% CI: 1.31 to 4.35). Women with inadequate GWG were therefore more likely to be multigravida. Male foetus conception was associated with increased odds of inadequate rate of GWG, AOR= 3.38 (95% CI: 1.05 to 10.91). The set of three predictors explained 31.4 % of the variance in inadequate GWG (Nagelkerke R Square =0 .314).

**Table 4:** Predictors of inadequate gestational weight gain per week (Logistic regressions)

|  | Wald | Significance | Adjusted odds ratio (AOR) | 95% confidence interval (CI) |  |
| --- | --- | --- | --- | --- | --- |
|  |  |  |  | Lower | Upper |
| First trimester BMI | 17.41 | <0.001 | 0.69 | 0.58 | 0.82 |
| Male foetus | 4.14 | 0.042 | 3.38 | 1.05 | 10.91 |
| Gravidity | 8.13 | 0.004 | 2.39 | 1.31 | 4.35 |
| Constant | 17.95 | <0.001 | 12717.48 |  |  |

### Relationship between inadequate GWG and length-for-age z score among infants under 6 months

In a multivariable linear regression, after adjusting for identified confounders, children of **wo**men with inadequate GWG had a significant lower length-for-age z scores (LAZ), compared with their colleagues who were born to women who had adequate GWG [beta coefficient (β), = -0.181 (95% CI: -2.72 to -0.14, p = 0.03)]. A unit increase in gravidity positively increased the mean LAZ by 0.23 standard units, [β = 0.23, (95% CI: 0.12 to 0.73, p = 0.006). A unit increase in infant’s birth weight increased LAZ by 0.27 standard units [β = 0.27 (95% CI: 1.15 to 4.71)]. Female infants had a mean LAZ that was significantly higher by 0.34 standard units (beta β = 0.34, p < 0.001), compared to male infants (Table 5).

**Table 5:** Predictors of LAZ (Multivariable regression analysis) among infants aged 0-5 months.

| Variable | Standardized Coefficients | t | Sig. | 95.0% Confidence Interval for $\beta$ | | Collinearity Statistics | |
| --- | --- | --- | --- | --- | --- | --- | --- |
| | Beta ($\beta$) | | | Lower Bound | Upper Bound | Tolerance | VIF |
| (Constant) |  | -4.05 | <0.001 | -16.24 | -5.58 |  |  |
| Gravidity | 0.23 | 2.80 | 0.006 | 0.12 | 0.73 | .981 | 1.019 |
| Inadequate GWG rate in grams per week | -0.18 | -2.20 | 0.030 | -2.72 | -0.14 | .942 | 1.061 |
| Birthweight | 0.27 | 3.27 | 0.001 | 1.15 | 4.71 | 0.955 | 1.047 |
| Female child | 0.34 | 4.18 | <0.001 | 0.91 | 2.56 | 0.984 | 1.016 |

Some potential confounders including age of child, first trimester BMI, mother’s occupation, maternal educational level, marital status, maternal age, ethnicity, obstetric and medical history of mothers during pregnancy, malarial infection during pregnancy were adjusted for but these were not significant and were therefore removed from the model. The set of variables that remained significant accounted for 25.3 % of the variance in LAZ (Adjusted R^2^ = 0.253)

### Relationship between rate of GWG and weight-for-age z score among children 6-24 months

Among children aged 6-24 months rate of GWG was negatively associated with weight-for-age z score but not height-for-age z score. In a multivariable linear regression, after adjusting for identified confounders, rate of GWG had a significant negative association with weight-for-age z score (WAZ), [beta coefficient (β), = -0.176 (95% CI: -0.006 to -0.001, p = 0.007)]. Children born to unemployed mothers had a mean weight-for-age z score which was -.141 standard units lower than their colleagues born to mothers employed in the Civil/Public Service (β = -0.141, 95% CI:-0.77 to -0.03, p = 0.03. A unit increase in dietary diversity score for children positively increased the mean WAZ by 0.234 standard units, [β = 0.234, (95% CI: 0.09 to 0.32, p = 0.001)]. Stunted children had lower WAZ than children not stunted (β = -0.133, (95% CI: -1.22 to -0.01). Children who were fed with colostrum at birth had higher WAZ compared to children who were not fed with colostrum [β = 0.159, 95% CI: 0.23 to 2.39, p = 0.02], Female children had a mean WAZ that was significantly higher by 0.18 standard units (beta β = 0.181, 95% CI: 0.17 to 1.08, p = 0.007), compared to male children (Table 6). The set of variables that remained significant accounted for 16.7 % of the variance in WAZ (Adjusted R^2^ = 0.167).

**Table 6:** Predictors of WAZ (Multivariable regression analysis) among children aged 6-24 months.

|  | Standardized Coefficients | T | Sig. | 95.0% Confidence Interval for (β) |  | Collinearity Statistics |  |
| --- | --- | --- | --- | --- | --- | --- | --- |
|  | Beta (β) |  |  | Lower Bound | Upper Bound | Tolerance | VIF |
| (Constant) |  | -3.306 | 0.001 | -6.587 | -1.664 |  |  |
| Unemployed Mothers | -0.141 | -2.137 | 0.03 | -0.77 | -0.03 | .959 | 1.043 |
| Rate of GWG | -0.176 | -2.713 | 0.007 | -0.006 | -0.001 | .993 | 1.007 |
| Stunted growth | -0.133 | -2.019 | 0.04 | -1.22 | -0.01 | .959 | 1.042 |
| Dietary diversity score for children | 0.234 | 3.470 | 0.001 | 0.09 | 0.32 | .915 | 1.093 |
| Female child | 0.181 | 2.723 | 0.007 | 0.17 | 1.08 | .945 | 1.058 |
| Child fed with colostrum | 0.159 | 2.393 | 0.02 | 0.23 | 2.39 | .939 | 1.065 |
| BMI of mother | 0.120 | 1.792 | 0.08 | -0.007 | .152 | .927 | 1.078 |

## Discussion

This study assessed the predictors of gestational weight gain and its association with post-natal growth of children under two years. The results showed that postnatal linear growth of children especially infants under 6 months is strongly dependent on adequate GWG rate. The main findings are discussed below:

### Prevalence of inadequate weight gain

A mean GWG of 162.0 g per week was observed in our study sample which is very low but consistent with what is reported from other low- and middle-income countries (LMICs) including Ghana [28, 29]. The majority (90.3%) had inadequate weight gain which also compares well with other findings in LMICs including Ghana [30-32].

The higher proportion of women with inadequate GWG in low-income sub-Saharan African settings may be due to a variety of nutritional problems, as well as frequent infections, poverty, and food insecurity [33]. Furthermore, a significant proportion of women in the Sub-Saharan region are not well informed on the risk factors associated with GWG [33]. Given the undesirable maternal and child health outcomes resulting from inadequate GWG [31], the findings of this current study further underscore the fact that inadequate GWG is a public health concern that warrants appropriate maternal health interventions during pregnancy to improve GWG.

### Predictors of inadequate GWG

A key finding in this study was that a high first trimester BMI protected against inadequate rate of GWG. A woman’s body mass index (BMI) during the first trimester was positively associated with her gestational weight gain (GWG) rate, which means that a higher BMI is associated with a higher GWG rate. This finding is consistent with previous studies conducted elsewhere [32, 34-39].

The positive association between first trimester BMI and GWG rate could be explained by the fact that women who are underweight at early pregnancy must gain more weight than their overweight or obese counterparts in order to achieve a healthy GWG. Women of higher BMI, on the other hand, would gain comparatively little weight to achieve adequate GWG as they are able to use some of their stored energy to support the growth of the foetus. As such, adequate GWG may be attained easily for such women.

Increased gravidity was associated with increased odds of low GWG, a finding which is consistent with earlier studies conducted in other countries including Niger and Tanzania [40, 41]. This association may partly be explained by the fact that higher gravidity may result in maternal nutritional depletion thereby affecting growth of the foetus and other tissues that contribute to GWG. Women who have been pregnant multiple times are at a greater risk of inadequate weight gain during pregnancy compared to women who are pregnant for the first time. Worth noting is the fact that multiple pregnancies may alter the body’s metabolic reactions or how the body stores and uses energy during pregnancy, which could have an impact on GWG. Additionally, multigravida might have more obligations including taking care of older children, which could limit their time or resources for attending to their own nutritional and weight-gain demands [42].

Male foetus conception was a risk factor associated with increased odds of inadequate rate of GWG. This implies sex of the developing foetus could have an impact on the mother’s weight gain during pregnancy. As shown by scientific studies, the energy balance, metabolism, and general GWG of mothers may be impacted by specific physiological and hormonal dynamics linked to male foetuses [43]. Pregnancy with a male foetus is associated with different hormonal patterns, such as slightly higher levels of testosterone. These hormonal variations may have an impact on GWG by altering the mother’s appetite, metabolism, or fat storage [44]. Higher testosterone levels, for example, may have an impact on insulin sensitivity, which may decrease GWG by influencing how well a mother’s body stores or uses energy. Male fetuses often have higher metabolic demands and may grow more quickly, especially in the later stages of pregnancy [45]. In order to promote an ideal foetal growth, the mother may need to consume extra calories. A woman’s GWG may, however, drop below the recommended levels if she does not increase her caloric intake to meet these increased needs. Some studies noted that women carrying male foetuses may experience different cravings or changes in appetite compared to those carrying female foetuses [44]. There may be a decrease in GWG if these changes result in reduced food intake or changes in dietary patterns.

We have noted that though some studies have identified age of mother, marital status, and educational status as significant determinants of gestational weight gain, these factors were not significant in the present study as has been reported in earlier studies [32, 34, 40].

### Association between GWG rate and infant postnatal growth

In this analytical cross-sectional study, after adjusting for potential confounders, inadequate GWG rate associated with lower length-for-age (LAZ) among infants aged 0-5 months. Similar findings have reported by some other investigators [46-49]. Children born to women with adequate GWG had significantly higher LAZ than those born to women who had inadequate GWG though this observation was not observed among older children aged 6-24 months. The lack of association beyond the early infancy period may be partly due to other more influential environmental factors including dietary intake and exposure to infections. Maternal nutritional status, which is a reflection of the amount of weight gain during pregnancy and energy stores can significantly impact foetal growth and early postnatal growth [50]. Infants, in this early growth of their life depend largely on their birth length and early nutritional status, which are strongly impacted by GWG and maternal health. An adequate GWG could leave the mother with enough energy stores for her foetus in-utero and its early life and vice versa. Similarly, during the first six months of life, weight and length growth are sharp. As a result, during this time, the impact of prenatal characteristics, such as GWG, are more noticeable. However, as the child grows beyond 6 months, other factors, such as postnatal nutrition, infections, and environmental conditions, start to play larger roles in influencing growth. Infants start to deviate from their birth trajectories as they adjust to their own growth patterns and environmental factors thereby lessening the impact of GWG.

The quality and quantity of food consumed by children post six months as mothers introduce complementary feeding could introduce a new nutritional dimension and significantly impact postnatal growth of older children. Abubakari et al. (2023), in a study conducted in the Tamale metropolis to determine the effects of maternal dietary habits and gestational weight gain on birth, also emphasized that dietary intake of an individual can be determined by culture and the type of food available to the person [51]. Therefore, this observation could account for the different growth patterns of children older than six months.

A unit increase in infant’s birth weight was associated positively with increased LAZ. This finding aligns with a retrospective cohort study that was conducted in the Northern Region of Ghana by Saaka and Abaah (2015), which also reported an association between LAZ and birth weight. This study further noted that the age of a child determines the effect of birth weight on LAZ. Children between the ages of 6-8 months who were born low birth weight tend to grow faster in length than children who were born with optimal weight. Nevertheless, children between the ages of 9-12 months born with normal weight grow faster than low birth weight babies (Saaka & Abaah, 2015). This relationship emphasizes the importance of birth weight as a key determinant of early growth outcomes. Better birth weight could reduce the risk of stunting in later life imply that infants who are born heavier typically have better linear growth, or length, in relation to their age. The association between LAZ and birth weight provides further evidence that obtaining a suitable birth weight may establish the foundation for normal growth patterns. Healthy birth weights can have a positive impact on children’s early linear growth, and interventions to enhance the nutrition and general health of the mother throughout pregnancy can help achieve this.

Female infants had a mean LAZ that was significantly higher, compared to male infants. This finding suggests that, on average, female infants tend to have better linear growth (or are less likely to experience stunting) than male infants. In general, male children are more likely than their female counterparts to suffer from stunting, especially in lower socioeconomic groups and Sub-Saharan Africa [52, 53]. This risk could be due to biological differences in growth rates, metabolism, or body composition between male and female infants in early life. According to some research, in some unfavorable circumstances, female babies may have a biological advantage that results in marginally better growth outcomes than boys [52]. This resilience could be the reason for which female infants usually achieve higher LAZ scores than males.

Among children aged 6-24 months, the rate of GWG was negatively associated with weight-for-age z score but not length-for-age z score. GWG is clearly about weight, so it is appropriate that it is associated with weight-related outcomes, but the negative association observed is rather unexpected. It appears lower GWG in foetal life may be compensated for during postnatal period such that children that did not gain enough weight during the foetal tend to gain weight faster in the postnatal period.

## Conclusion

The study has identified first trimester BMI, conception of male foetus and gravidity to be significant predictors of GWG. The association between GWG and postnatal growth outcomes suggests that targeted interventions are needed to address maternal nutrition, particularly among multigravida women and those with lower pre-pregnancy BMI. It is, therefore, recommended that interventions should focus on ways to improve healthy GWG so as to reduce the prevalence of child stunting.

## Strengths and limitations of the study

This study is one of the few in lower-income countries and the first in Ghana to assess the association of rate of gestational weight gain on postnatal growth outcomes. Despite its strengths, this study has several limitations. First, the present study was cross-sectional and therefore causality cannot be inferred. The classification of GWG rate was based on the recommendations by US Institute of Health which may underestimate the proportion of gestational weight gain in developing countries such as Ghana, where this study was conducted. Furthermore, early pregnancy BMI was taken before or at 12 weeks of gestation, at which time there may already have been an increase or decrease of gestational weight.

## Authors’ contributions

MS and AMD conceived the study, participated in its design and contributed significantly to the acquisition of data. MS did the analysis and interpretation of data and together with AMD drafted the manuscript and revised it critically for important intellectual content. Both the authors read and approved the final draft.

## Acknowledgements

The authors wish to acknowledge with gratitude the contribution of the data collection team whose hard work and commitment led to a successful conduct of this study. The co-operation and support of mothers and caregivers who took time off from their busy schedules to respond to the interviewers is very much appreciated.

## Data Availability

The quantitative data presented in the form of SPSS used to support the findings of this study are included within the supplementary information file(s).

